# Inferring variant-specific effective reproduction numbers from combined case and sequencing data

**DOI:** 10.1101/2021.12.09.21267544

**Authors:** Marlin D. Figgins, Trevor Bedford

## Abstract

Accurately estimating relative transmission rates of SARS-CoV-2 variants remains a scientific and public health priority. Recent studies have used the sample proportions of different variants from genetic sequence data to describe variant frequency dynamics and relative transmission rates, but frequencies alone cannot capture the rich epidemiological behavior of SARS-CoV-2. Here, we extend methods for inferring the effective reproduction number of an epidemic using confirmed case data to jointly estimate variant-specific effective reproduction numbers and frequencies of co-circulating variants using cases and sequences across states in the US from January 2021 to March 2022. Our method can be used to infer structured relationships between effective reproduction numbers across time series allowing us to estimate fixed variant-specific growth advantages. We use this model to estimate the effective reproduction number of SARS-CoV-2 Variants of Concern and Variants of Interest in the United States and estimate consistent growth advantages of particular variants across different locations.

## Introduction

As SARS-CoV-2 evolves, variants may emerge that increase in their ability to transmit and escape acquired immunity [1]. Quantifying the observed growth advantages of SARS-CoV-2 variants allows us to better understand biological differences between circulating viruses [2, 3]. Relating genomic data of SARS-CoV-2 variants to epidemic surveillance data is difficult. Although it is typical to use phylodynamic methods to analyze genetic sequence data from epidemics, the sheer amount of data as well as challenges to describing fitness effects in phylodynamic models make these methods hard to apply to potential differences in transmission rate among circulating variants. In order to deal with the limitations of phylodynamic inference, previous studies have estimated the growth of variants using observed frequencies in sequenced SARS-CoV-2 samples [4–7]. Such methods often model the frequency of variants using multinomial logistic regression [4, 6], which generally assumes that genetic variants have a fitness advantage over one another which is fixed in time and acts as a estimate for the selective advantage of different variants at the level of frequencies. Although a consistent increase in frequency of one variant over another is expected to reflect differences in transmission rate, these models do not directly account for the complicated infection and transmission dynamics which influence which variants lead to local and regional epidemics. When dealing with competition between variants, variants which are declining in frequency can still lead to an increasing number of infections. Similarly, growth in frequency does not necessarily entail an increase in absolute infections.

To more fully capture epidemiological dynamics, there are methods which describe the growth in number of infections using confirmed case, hospitalization, or death data to estimate changes in the effective reproduction number *R*_*t*_, the average number of infections a single infectious individual generates at a given point of time *t*. Although these methods are excellent for describing overall epidemic growth rates, they cannot capture the evolutionary dynamics and fitness changes between different variants since they generally assume the population dynamics are described by a singular *R*_*t*_ trajectory [8, 9], which internally is unrelated to the genetic and phenotypic composition of the population. This is of particular importance in the analysis of an epidemic in which a dominant variant may be declining overall, but a minor variant is rapidly increasing in frequency and absolute prevalence, creating the potential for a secondary wave of infections that may go unnoticed at first glance. To overcome this we require models that partition case counts into contributions from different variants to estimate variant-specific effective reproduction numbers.

Ongoing SARS-CoV-2 evolution serves as an important example of this phenomenon. After initial emergence in late 2020, over the course of 2021, Variant of Concern (VOC) and Variant of Interest (VOI) viruses spread throughout the world and replaced existing viral diversity. Multiple WHO designated [10] VOC and VOI viruses circulated in spring and early summer 2021, but this diversity was largely replaced by Delta variant viruses which became globally dominant in late summer 2021. Subsequently, Delta variant viruses were rapidly eclipsed by Omicron variant viruses after Omicron’s emergence in October 2021 [11]. Although it is now clear that Delta’s spread was driven by greater transmissibility than other co-circulating variants and Omicron’s rapid spread was primarily driven by escape from existing population immunity, rigorous estimates of the relative fitness of circulating variant viruses are of interest. Here, we develop a joint epidemiological and population genetic model of SARS-CoV-2 to assess the growth of different variants over time and infer differences in the effective reproduction numbers of SARS-CoV-2 variants as well as underlying frequency of variants under noisy sampling. We apply this model to sequence data and case count data from the United States between January 2021 and March 2022 to estimate differences in transmissibility between circulating VOC and VOI viruses.

## Results

### Model Overview

We implement two models of variant-specific effective reproduction number based on a renewal equation framework of epidemic spread (see Methods), a fixed growth advantage model and a time-varying growth advantage model (growth advantage random walk — GARW). These models assume that new infections are determined by two essential parameters: the effective reproduction number which determines the average number of secondary infections generated over the course of a primary infection and the generation time which determines length of infection and relative transmissibility over the course of the infection. In both models, variants generate infections independently of one another, but the sum of infections across variants is observed through surveillance data like case counts or hospitalizations. In order to disaggregate infections by variant we rely on frequency estimates which are informed by counts of sequenced samples using a Dirichlet-multinomial likelihood.

The transmission of each variant is modeled using a deterministic renewal equation which allows for realistic delay distributions between infection, transmission, and detection as a case. With this approach, we need only to determine the initial number of infections and the variant-specific effective reproduction numbers to estimate the frequency of each variant in the population over time. Due to this, the differences between the two models is determined in how each parameterizes variant-specific effective reproduction numbers.

Each variant in the fixed growth advantage model has its own multiplicative growth advantage which acts as a scaling to a single non-variant *R*_*t*_ trajectory (Fig. 1). With this fixed growth advantage model, we parameterize fitness of variants at the level of transmission by inferring variant-specific effective reproduction numbers. This differs from previous work on variant effective reproduction numbers which often parameterize these differences by assuming logistic growth of frequencies [12, 13]. Though, in general, our method allows one to estimate variant growth in the frequency domain in terms of effective reproduction number differences, we find that assuming a fixed advantage for variants results in estimates which are qualitatively similar to the aforementioned models which assume fixed growth advantages in frequency growth. This model provides the benefit of the inferred parameters being interpretable as scaling the effective reproduction number.

**Figure 1.**
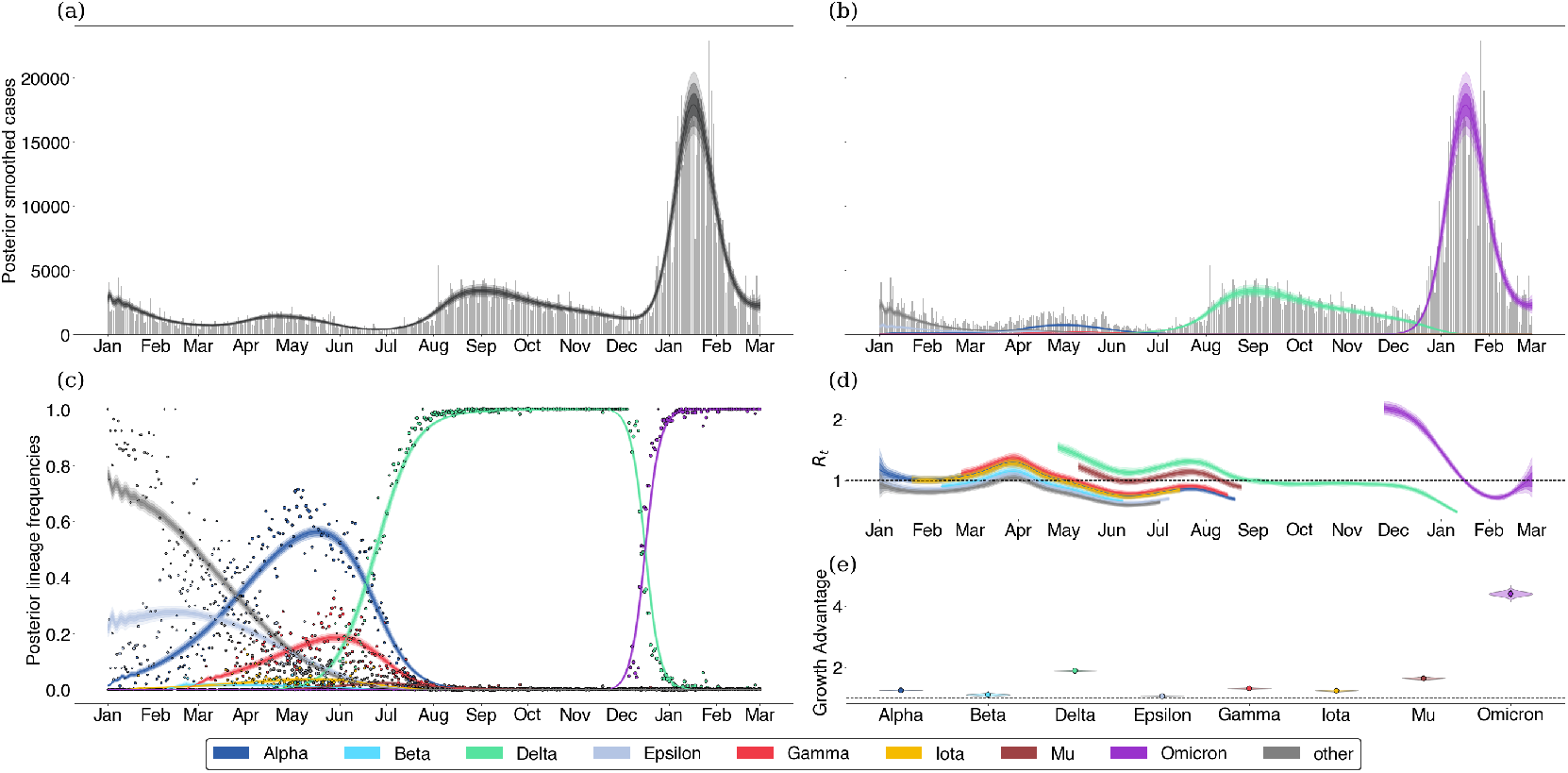
Fitting the fixed growth advantage model to Washington state data. (a) Posterior expected cases without weekly seasonality in reporting rate. Gray bars are observed daily case counts, while blue lines are model inferences with 50%, 80% and 95% credible intervals. (b) Posterior expected cases by variant. Each colored line is a different variant with intervals of varying opacity showing 50%, 80% and 95% credible intervals. (c) Posterior variant frequency against observed sample frequency. Dots represent observed weekly frequencies in sequence data and each colored line is a different variant with shaded CIs. (d) Variant-specific effective reproduction numbers. (e) Posterior growth advantage by variant.

In cases where a singular fixed growth advantage is insufficient to describe the data, we extend our model to allow time-varying growth advantages (Fig. 2). In the GARW model, we introduce a variant *R*_*t*_ which infers the effective reproduction number of each variant as having a time-varying growth advantage relative to a base variant to allow for more complex relationships between the growth rates of different variants over time. Each variant effective reproduction number is parameterized using an exponentiated spline basis, so that the log effective reproduction numbers are described by a linear basis expansion. Therefore, we can use smoothing priors on the coefficients of these basis expansions to regularize the inferred time-varying growth advantages of each variant.

**Figure 2.**
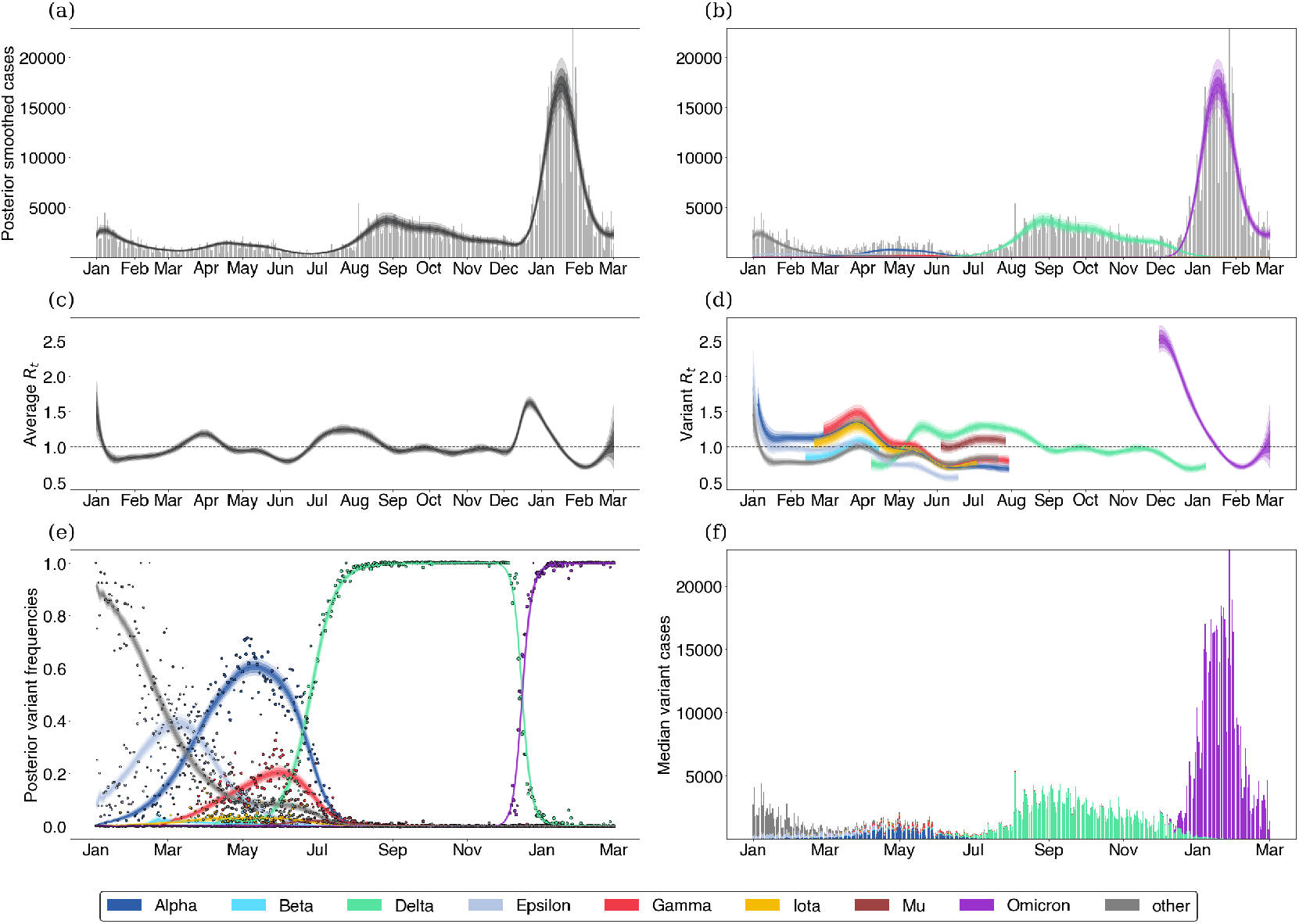
Fitting the GARW model to Washington state data. (a) When assessing epidemic growth rates, we often compute a single effective reproduction number trajectory which is effectively an average over the all viruses in population. We show the posterior smoothed incidence over time as well as the average effective reproduction number. Gray bars are observed daily case counts, while black intervals are the posterior 50%, 80% and 95% credible intervals. (b-d) Epidemics are made of different variants which may differ in fitness. We show the posterior variant-specific smoothed incidence (b) as well as the average and variant-specific effective reproduction numbers (c-d). (e-f) Using case counts alongside sequences of different variants allows us to understand the proportion of different variants in the infected population.

We demonstrate these models on data from Washington State with results from the fixed growth advantage model shown in Figure 1 and results from the GARW model is shown in Figure 2. Further example model output for California, Florida, Michigan and New York is provided in the supplemental appendix in Figures S1–S8.

### Estimating growth advantages in the United States

We estimate the effective reproduction numbers of SARS-CoV-2 Variant of Concern and Variant of Interest viruses in the United States using daily confirmed case counts obtained from the US CDC and sequence counts annotated by variant obtained from the Nextstrain-curated ‘open’ dataset [14] (see Data and code accessibility). Each sequence is labeled with a Nextstrain clade [14], and we partition clades into variants based on WHO VOC/VOI designation [10]. Nextstrain clades annotated in the fashion correspond to a subset of lineages designated by Pango [15]. We consider the following 8 variants which have been flagged as variants of interest or concern and which circulated in the US during 2021 and early 2022: Alpha (Pango lineage B.1.1.7, Nextstrain clade 20I), Beta (lineage B.1.351, clade 20H), Gamma (lineage P.1, clade 20J), Delta (lineage B.1.617.2, clade 21A), Epsilon (lineage B.1.427/429, clade 21C), Iota (lineage B.1.526, clade 21F), Mu (lineage B.1.621, clade 21H) and Omicron (lineage B.1.1.529, clade 21M). We use a cutoff of 2000 sequences from a particular variant across states to determine threshold of circulation. This eliminates Eta, Theta, Kappa and Lambda from consideration and groups these variants along with ancestral ‘non-variant’ viruses into a single ‘other’ category. We use a cutoff of 12,000 sequences from a particular state as basis for including the state in the dataset. This cutoff left 34 states available for inference.

In order to inform our estimates of the frequency of genetic variants, we divide sequences from each state into daily sample counts for each of the 8 variants above and a single ‘other’ category. We then use these counts alongside the daily case counts in each state to estimate the effective reproduction number for individual variants using the GARW *R*_*t*_ model. We find that overall there appears to be consistent trends in the effective reproduction numbers of variants across the United States (Fig. 3). We see that early VOCs Alpha and Gamma initially had *R*_*t*_ > 1, but saw *R*_*t*_ decline below one across most states in April and May respectively. Upon arrival in May, Delta shows significantly higher values of *R*_*t*_ that don’t decline below 1 until September. Initial Omicron *R*_*t*_ in November and December is significantly greater than earlier variants, but declines below 1 in late January and early February after driving large epidemics across states.

**Figure 3.**
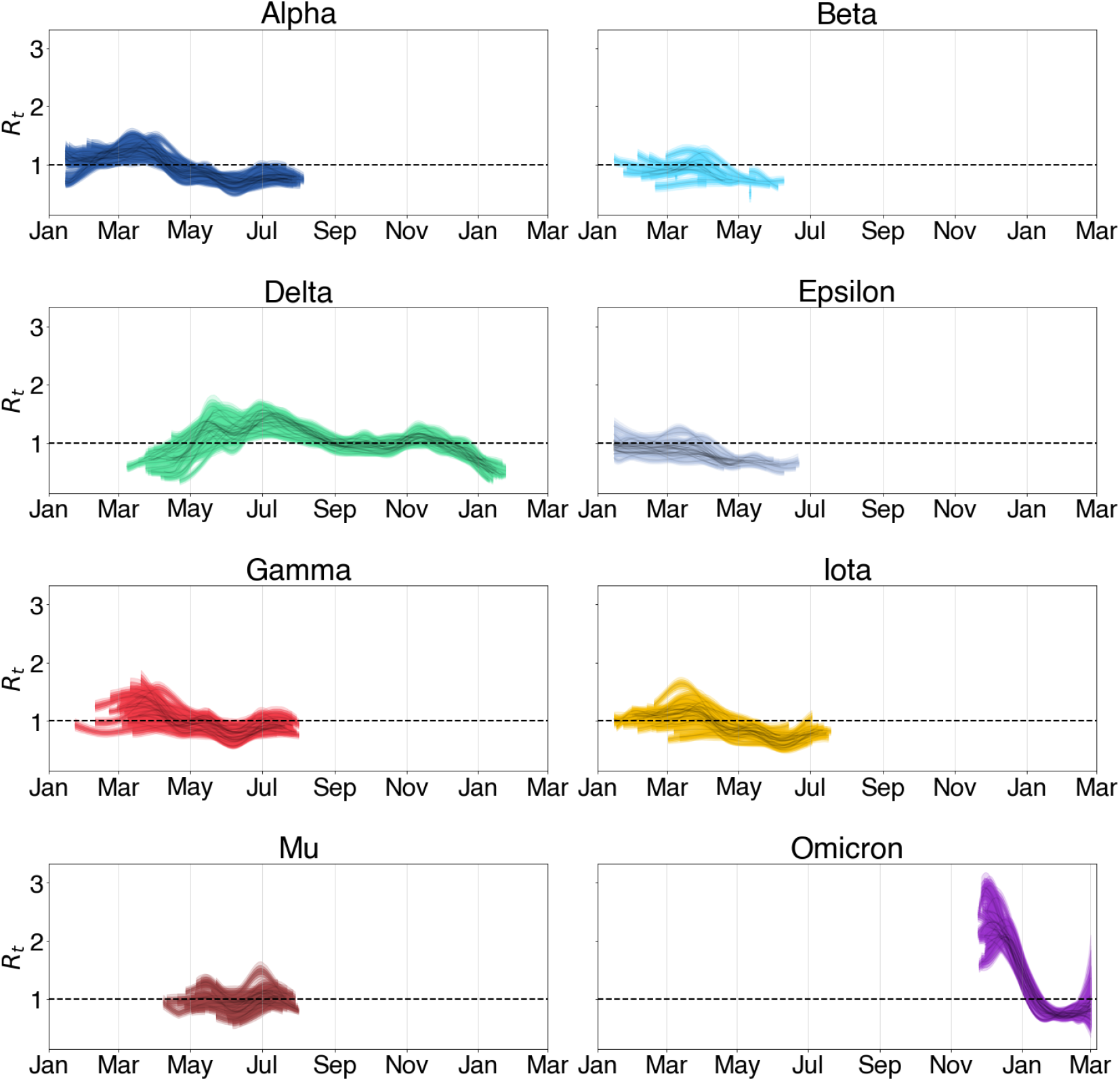
Inferred effective reproduction numbers from GARW model in 34 states show consistent trends of variants across states. Each panel shows a series of 34 trajectories, representing *R*_*t*_ through time for this variant across states. Shaded intervals show 50%, 80% and 95% credible intervals.

In order to transform these observed trends to a variant-specific growth advantage, we rely on our fixed growth advantage model which infers a fixed variant-specific growth advantage as a multiplicative scaling of the effective reproduction number. Using the fixed growth advantage model, we find that most variants identified share some positive growth advantage except for Epsilon (Fig. 4). Further, these growth advantages appear to be consistent between the states analyzed. These results from the fixed growth advantage model are consistent with a multinomial logistic growth analysis (Fig. S9). Alpha, Beta, Gamma and Iota show modest growth advantage over largely ancestral ‘other’ viruses, while Mu and Delta show larger growth advantages. Mu has previously been associated with increased neutralization resistance to convalescent serum [16], and its advantage of 1.2–1.8 across states is perhaps partially driven by immune escape. Despite this, Mu’s growth advantage whether from immune escape or otherwise was insufficient to outcompete Delta in any of the states analyzed. Delta’s advantage of 1.6–2.0 across states is particularly significant. Given this large growth advantage was evident in May (Fig. 3), Delta’s rapid rise in frequency and sizable epidemic should have been clear at the time. The significant growth advantage observed in Delta is recapitulated in other studies including Obermeyer et al. [6] and Vöhringer et al. [17]. In the case of Omicron, we see significant variability in the growth advantage which spans 2.0–4.4. This large variability could be motivated by multiple factors including state-level variation in population immunity.

**Figure 4.**
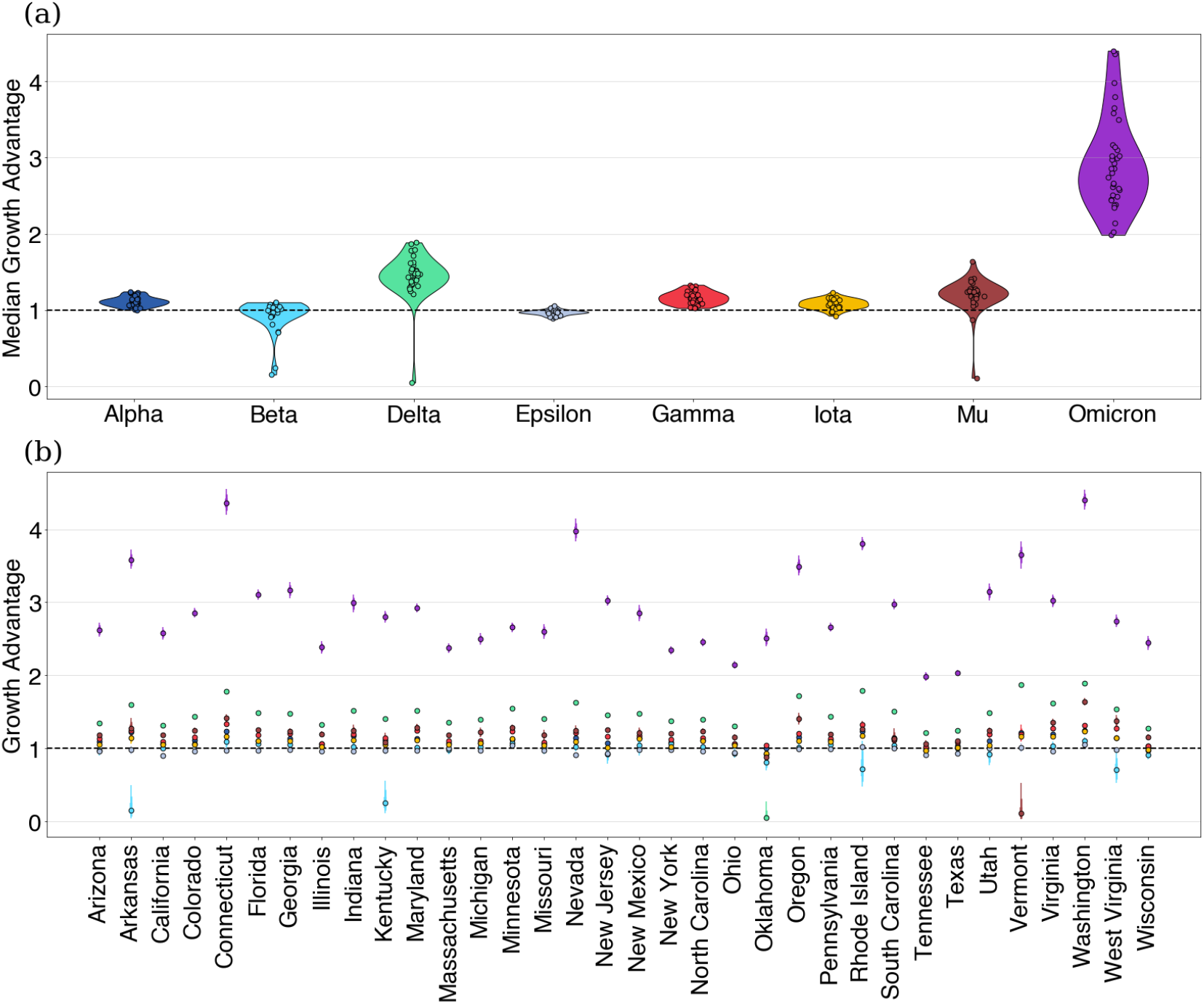
Using fixed growth advantage model, we infer growth advantages for 8 variants in 34 US states. (a) Growth advantages for variants of concern. Each point is the median growth advantage inferred from a single state. (b) Same as (a) but with state medians visualized by variant.

To better address the potential for change in variant growth advantage over time, we use our GARW model on the same data set to assess how variants increased or decreased in their growth advantages over time. We see growth advantages which are overall consistent with our fixed growth advantages, but are clearly able to discern time periods of variable growth advantage (Fig. 5). We observe oscillations in Delta’s growth advantage from June to January. For Omicron, we also observe a large variability in the time-varying growth advantage though there appears to be an upward trend after December.

**Figure 5.**
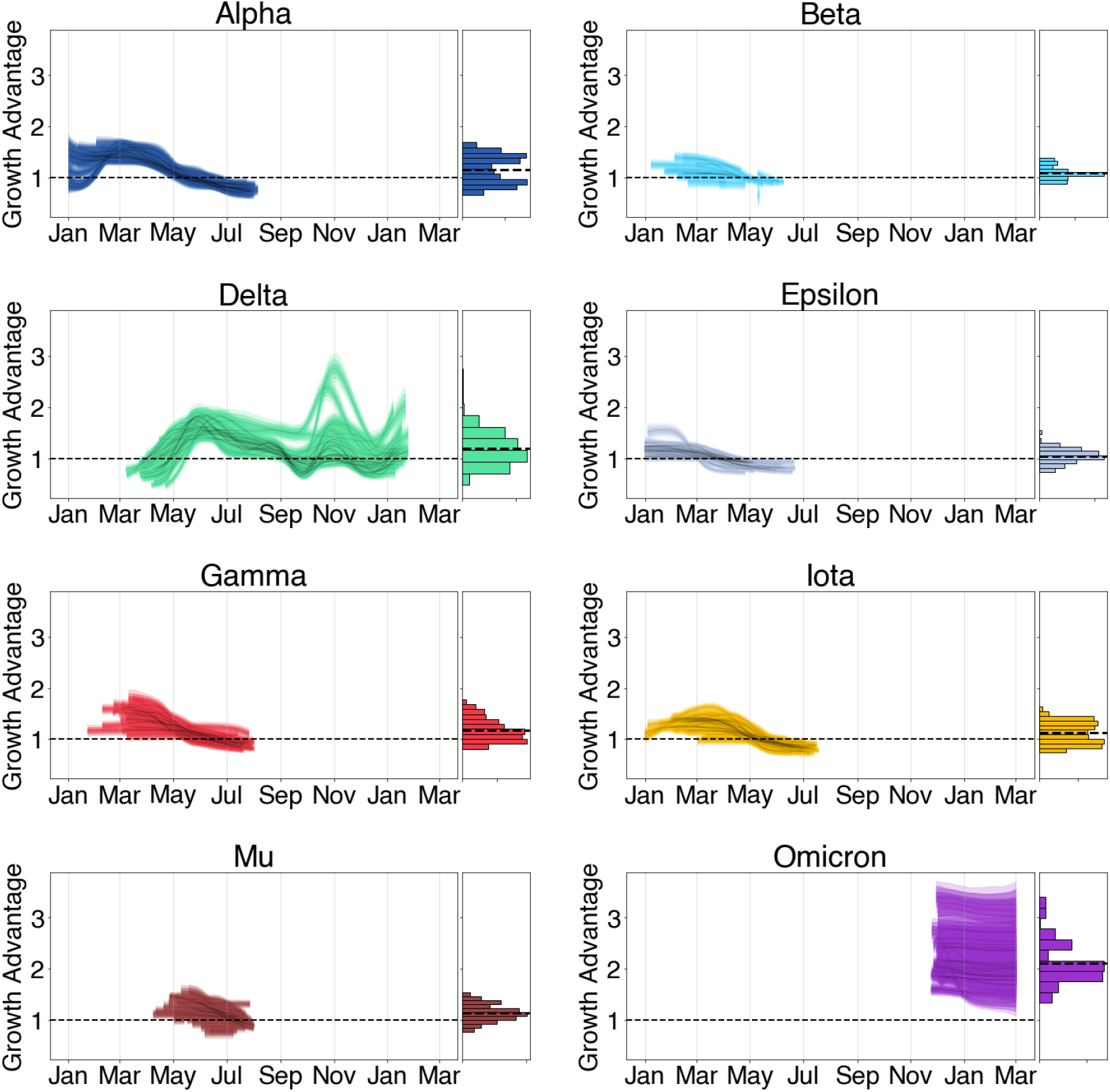
Estimating variant growth advantages in 34 states using GARW model. Each panel shows a series of 34 trajectories, representing Δ_*v*_ through time for variants across states. Histograms show the distribution of the variant’s growth advantage over time. Shaded intervals show 50%, 80% and 95% credible intervals.

## Discussion

We find that a model that partitions case count data based on variant frequency in sequence data works well to describe SARS-CoV-2 variant dynamics in the United States from January 2021 to March 2022. In each state, spring waves in 2021 were primarily driven by the arrival of Alpha, Beta, Gamma, and Iota variants. However, as these waves subsided, the arrival of Delta with a significantly greater growth advantage, drove a large wave in summer 2021. Omicron’s arrival in November 2021 drove a much larger wave in January/February 2022 due to significant immune escape of the variant. Importantly, we can directly estimate a variant-specific *R*_*t*_, which for example, shows that Delta was a growing rapidly sub-epidemic across states in May, before its impact was noticeable in overall case counts, and that Omicron’s initial *R*_*t*_ was estimated to be between 2 and 3 in December 2021, presaging a substantial Omicron driven wave.

We imagine that this approach could provide early warning of imminent epidemics driven by low-frequency but highly transmissible variants and generally serve to identify newly arising variants that show significant transmission advantages and that may drive epidemics. Indeed, we have continually updated estimates of spread of Omicron and Omicron sublineages BA.2, BA.2.12.1, BA.4 and BA.5 using this method and shared results in realtime online at github.com/blab/rt-from-frequency-dynamics. As an example, we estimate the growth advantages of Omicron sublineages BA.2, BA.2.12.1, BA.4, and BA.5 during their rise in the United States in Figure 6. These real-time estimates have served as a basis for reporting to public health, policy makers and the general public.

**Figure 6.**
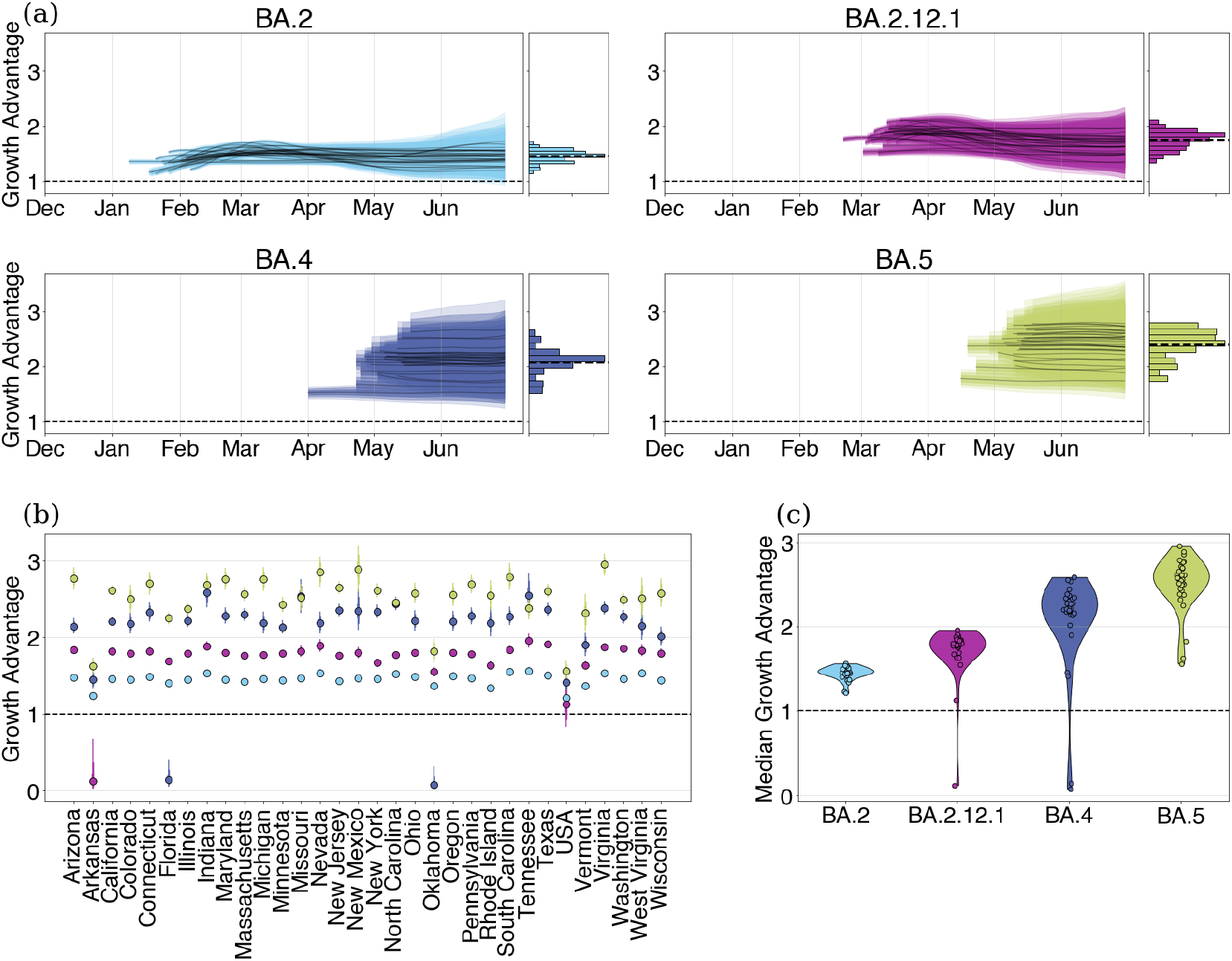
Estimating growth advantages of Omicron sublineages relative to BA.1 in 33 US states. (a) Time-varying growth advantages for BA.2, BA.2.12.1, BA.4, and BA.5 relative to BA.1 using the GARW model. Histograms denote the distribution of the variant growth advantages across all times. (b) Fixed growth advantages for Delta and BA.2 relative to BA.1 using fixed growth advantage model. (c) Same as (b) but with state medians visualized by variant.

With this mind, this work is not without limitations. The underlying transmission model is deterministic and does not account for demographic stochasticity and over-dispersion in transmission which has been documented in SARS-CoV-2 transmission [18]. As with all methods which depend on parameterizations of the generation time, misspecification of the generation time can be lead to biased estimates of the effective reproduction number or growth advantages [19]. In order to quantify this source of error, we derive an equation relating our inferred growth advantages, the epidemic growth rates, and the mean and standard deviation of the generation time distribution. This source of error can be partially combated by converting effective reproduction numbers to their corresponding epidemic growth rates under the generation time assumption (see Supplement Appendix). There is also a general need to account for biases in the case data which may not faithfully describe the infection dynamics of SARS-CoV-2 due to changes in case ascertainment rate, as possibly caused by differences in testing intensity, infection severity among other reasons. However, we suspect that case ascertainment remained largely consistent from January to ∼Dec 2021, even if it declined with the advent of widespread circulation of Omicron.

We do not explicitly model additional introductions of variants outside a fixed seeding period which can play an important role in variants establishing themselves in different geographies at low infection counts and could bias our estimates of the effective reproduction number if not properly accounted for [8, 20]. However, we expect once local transmission is predominant that estimated *R*_*t*_ will reflect characteristics intrinsic to the variant in the local geography. Using hierarchical models of variants to jointly estimate growth advantages and pool estimates across locations could be a useful approach for analyzing consistency between growth advantages of variants geographically and beginning to combat the issue of multiple introduction events. That said, fully combating this issue would likely involve incorporating demographic stochastic into the model at the level of transmission and likely reduce the speed of inference, scalability, and limit available inference options.

Although there are several ways to improve these methods and expand their applicability, our current model does have utility as a way of assessing early claims of variant advantages and is able to show there is evidence of consistent variant advantages shared between different geographies. Additional work is needed to attribute these inferred advantages to biological mechanisms like immune escape and transmissibility [1]. Modeling the effect of changes in other factors such as contact patterns or non-pharmaceutical interventions can be done with the current formulation of the model by including quantities of interest as features in the *R*_*t*_ model as in Sharma et al. [21].

In general, the development of methods which can account for fitness differences between genetic variants is much needed in order for proper epidemic preparedness. Our method provides one way of analyzing the growth rates of SARS-CoV-2 variants without directly parameterizing how variants grow in terms of frequency by instead focusing on differences in the effective reproduction number. In cases where the assumption of a fixed growth advantage is warranted and justified, our fixed growth advantage model provides a way of quantifying variant growth advantages at the level of transmission which allow for various delays between infection, transmission, and sampling. When a fixed growth advantage is unjustified, our GARW model can be used infer trends in variant growth advantages over time. Currently, our GARW model can be used to assess claims of growth advantages of variants and their sublineages.

This method can be extended further to analyze the role of specific constituent mutations defining a variant or lineage in changing the effective reproduction number of specific variants directly, similar to the model formulation of Obermeyer et al. [6]. With this in mind, our method potentially has use for evolutionary forecasting of variants for SARS-CoV-2 as we inform the frequency dynamics of co-circulating variants by describing their population-level transmission dynamics [22]. Extending the model further towards this aim will likely require methods for quantifying various sources population immunity as well as escape potential for circulating and emerging SARS-CoV-2 variants as a way to explain these growth advantages and their underlying mechanisms using data. With these issues in mind, surveillance of variants should be folded into standard epidemiological surveillance as knowledge of variant-specific growth advantages will be useful for forecasting growth of cases, hospitalization, deaths, vaccine effectiveness among other key metrics related to epidemic response.

Further, as case surveillance for COVID-19 has decreased in reliability after 2022, we note that this method is still applicable using other proxies for infection incidence such as hospitalization data or wastewater testing. However, even in the absence of these forms of data, our approach highlights the distinction between relative fitness of viral variants and their overall transmission rates allowing us to attribute changes in incidence to selection and variant turnover.

## Methods

Using sampled counts of sequences from different variants as well as case data, we can jointly infer the proportion of variants in the larger population and the effective reproduction number of these variants.

### Modeling the infection process

We estimate the effective reproduction number of competing variants using a deterministic renewal equation based framework. These equations arise as the expectation of a Bellman-Harris branching process [23] which is a type of Branching process in which offspring generation depends on the age of infection.

The renewal equation framework allows one to model infection processes in a way that is mathematically equivalent to standard epidemic models like the SEIR compartment model [24], but in a way that can be more suitable for estimating the effective reproduction number and forecasting using arbitrary generation times. This renewal equation can be written as

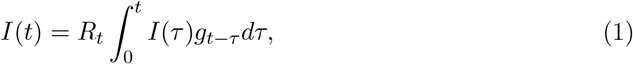

where *g* is the generation time. In addition, we also include onset distribution *o* for symptoms which allows us to compute the prevalence, or the number of active infections, as

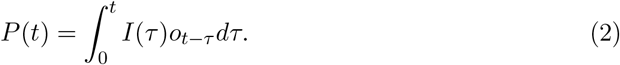

We bin the generation time *g* and the onset distribution *o* to the nearest day, so that we estimate the daily incidence *I*(*t*) and prevalence *P* (*t*) as

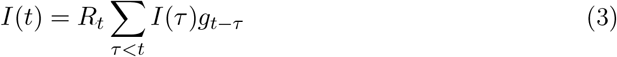

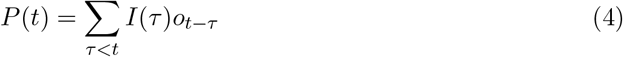

We parameterize all variants excluding Delta and Omicron as having generation time *g* as having Gamma distribution with mean 5.2 and standard deviation 1.2 in line with the estimates of [25]. Due to observed shorter serial intervals for Delta and Omicron, we instead use a mean generation time of 3.6 for Delta and a mean of 3.2 for Omicron [26–28]. For all variants, we parameterize the onset time *o* as having LogNormal with mean 6.8 and standard deviation 2.0 in line with [29]. We note that the choice of generation time can have strong effects on the inferred effective reproduction number and growth advantage under renewal equation model. The effect of generation time choice is quantifiable as shown in Figures S10, S12 and Supplemental Appendix (see Relating epidemic growth rates to relative effective reproduction numbers). Though converting the posterior effective reproduction numbers to epidemic growth rates may be more robust to changes in generation time as can be seen in Figure S11.

This method of using delays to represent lags between infection and observation can be extended to use multiple delays to better fit other data sources such as hospitalization or deaths.

### Modeling variant frequencies

In the case of *V* variants co-circulating in a population, we denote incidence of variant *v* at time *t* as *I*_*v*_(*t*) and prevalence as *P*_*v*_(*t*). In this case, we can compute the frequency of variant *v* in the population at time *t* under the infection process outlined above as

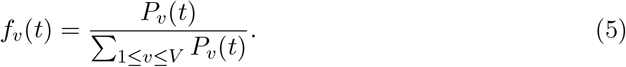

Since we’ve defined the frequency in terms of the transmission dynamics, the variant-specific effective reproduction numbers *R*_*t,v*_ and initial infections *I*_*v*_(0) determine the frequency dynamics directly. Therefore, we do not need to impose a parametric form on *f*_*v*_(*t*) directly as in other models of variant frequency.

### Observation process for cases

As most case time series in the United States and elsewhere exhibit day of the week biases, we estimate a reporting rate which varies by day of the week, so that *ρ* = (*ρ*_1_, …, *ρ*_7_) as in [9]. We then define the observation likelihood using a negative binomial distribution as follows

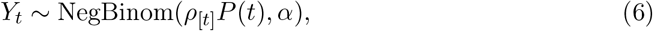

where [*t*] = *t* mod 7 +1, *α* is an over-dispersion parameter relative to the Poisson distribution and NegBinom(*µ, α*) is the negative binomial distribution with mean *µ* and variance *µ* + *αµ*^2^. In the case of multiple variants, we use *P* (*t*) = Σ_1≤*v*≤*V*_ *P*_*v*_(*t*). The negative binomial likelihood is often used for modeling observation noise for count data such as epidemic time series which are often over-dispersed relative to a Poisson distribution. In order to account for zero-counts due to a lack of observations, we also include zero-inflation on the case counts.

### Observation process for variant annotations

Suppose we’re tracking the growth of *V* variants, our data for a given day *t* takes the form of daily counts *C*_*t*_ = (*C*_*t*,1_, …, *C*_*t,V*_) of sequences of each variant with daily total *N*_*t*_ = Σ_1≤*v*≤*V*_ *C*_*t,v*_. We then assume that the likelihood of observing these counts of each variant is described by a Dirichlet-multinomial distribution, so that

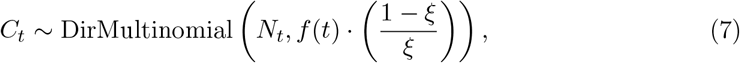

given variant frequencies *f* (*t*) = (*f*_1_(*t*), …, *f*_*V*_ (*t*)) and over-dispersion parameter 0 < *ξ* < 1. Here, we use a Dirichlet-multinomial distribution to account for possible over-dispersion in the counts relative to the standard Multinomial distribution.

### Basis expansions of log effective reproduction numbers

Instead of inferring *R*_*t*_ directly, we parameterize the log effective reproduction number using a basis of cubic splines. Each basis spline is written as a column in the design matrix ***X***, so that

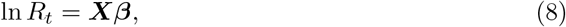

where the ***β*** are to be estimated to parameterize the effective reproduction number. We then use locally adaptive smoothing of order one with a Laplace prior on the coefficients ***β*** to promote smoothness on the inferred *R*_*t*_ trajectory [30]. This method also allows one to use other predictors such as vaccination proportion, intervention indicators, temperature, humidity, etc.

### Modeling variant-specific effective reproduction numbers

To model the variant-specific reproduction numbers, we can infer individual independent effective reproduction number trajectories for each variant

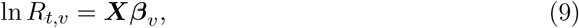

where each variant *v* gets its own vector of parameters ***β***_*v*_ in this model. We use the same prior structure as above to promote smoothness on inferred trajectories.

### Modeling variant-specific growth advantages

In order to use our model to infer growth advantages for specific variants, we can instead parameterize the effective reproduction numbers as

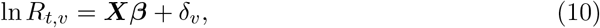

where the parameters *β* are shared between all variants and *δ*_*v*_ is the log-scale variant-specific growth advantage of variant *v*. We consider Δ_*v*_ = exp(*δ*_*v*_) to be the variant-specific growth advantage which can be seen in Figure 4. This model is referred to as the “fixed growth advantage model” throughout the paper.

### Estimating time varying growth advantages

In reality, the growth advantage of a variant may vary in time due to factors like cross-immunity between variants, overall immune escape, etc. This can additionally occur under variant generation time misspecification [31].

To combat these issues, we extend our model to allow for time-varying growth advantages. We consider a growth advantage random walk model (GARW) in which the time-varying variant growth advantage *δ*_*t,v*_ relative to a chosen “base” variant is modeled as a spline whose coefficients ***β***_*v*_ have a Laplace Random Walk prior

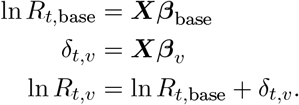

This model is referred to as the GARW model throughout the paper and can be seen in Figure 2

### Estimating an average effective reproduction number for an epidemic

Given variant-specific effective reproduction numbers *R*_*t,v*_ and the frequency of variants in the population *f*_*v*_(*t*), we define the average effective reproduction number to be

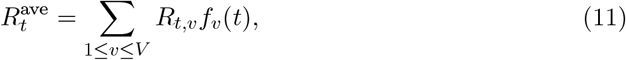

which is the sum of the variant-specific effective reproduction numbers weighted by their frequency. This quantity can be seen in Figure 2.

### Priors for Bayesian Inference

For both models, we provide a Laplace random walk prior on the spline coefficients ***β*** with scale parameter *γ* which itself has a HalfNormal(0, 0.1) prior distribution. In the fixed growth advantage model, only a baseline *R*_*t*_ trajectory is parameterized by ***β*** and the variant advantages *δ*_*v*_ are given a Normal(0, 1) prior. For the GARW model, the variant growth advantage spline coefficients are modeled with a Laplace random walk with scale parameter *γ*_*δ*_ which has HalfNormal(0, 0.01) prior distribution. The initial infected individuals for each variant have a uniform prior between 0 and 300,000. The weekly reporting rates *ρ*_[*t*]_ each follow a Beta(5, 5) prior, and the case observation over-dispersion is given a HalfNormal(0, 0.05) prior on 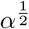. Finally, the over-dispersion parameter *ξ* is given a Beta(1, 99) prior to penalize high levels of over-dispersion in sequencing.

### Inference

The model is implemented in NumPyro [32] in Python and approximate Bayesian inference was conducted using Stochastic Variational Inference [33] using the ADAM optimizer [34] with a learning rate of 0.01. For the analyses presented, all models are fit using a full-rank Gaussian variational distribution / Multivariate Normal autoguide as implemented in NumPyro [32] which approximates the posterior (with appropriate constraints on the individual parameter spaces) as a multivariate normal distribution.

Models for each individual state in the United States variants data set were fit for 60,000 iterations and 3000 posterior samples were produced under both the fixed growth advantage model and the GARW model.

## Data Availability

Derived data of sequence counts and case counts, along with all source code used to analyze this data and produce figures is available via the GitHub repository \href{https://github.com/blab/rt-from-frequency-dynamics/}{github.com/blab/rt-from-frequency-dynamics}.

https://github.com/blab/rt-from-frequency-dynamics/

## Data and code accessibility

Case counts and sequence data was obtained March 26, 2022. Case count data was obtained from the US CDC using the ‘United States COVID-19 Cases and Deaths by State over Time’ dataset available from data.cdc.gov. Sequence data including date and location of collection as well as clade annotation was obtained via the Nextstrain-curated ‘open’ dataset [14] that pulls from sequences shared to NCBI GenBank. Sequence metadata is available from data.nextstrain.org. Clades in this dataset are assigned via Nextclade annotation [35]. Here, we subsetted to sequences with specimens collected from the USA between January 1, 2021 and March 1, 2022. We additionally filtered to sequences with known collection date, assigned Nextstrain clade and dropped samples that were flagged as ‘bad’ by Nextclade QC. This subsetting resulted in 1,906,759 sequences for analysis. However, we reduced dataset to just the 34 states with more 12,000 sequences available in this timeframe. Doing so reduced the full dataset to 1,541,099 sequences for analysis.

Derived data of sequence counts and case counts, along with all source code used to analyze this data and produce figures is available via the GitHub repository github.com/blab/rt-from-frequency-dynamics.

## Author contributions

MF, TB conceived the study. TB gathered sequence and case count data. MF designed and implemented inference model. MF performed the analysis. MF, TB interpreted the results. MF, TB wrote the paper.

## Acknowledgements

We thank John Huddleston, Eslam Abousamra and other members of the Bedford Lab for helpful feedback. MF is an ARCS Foundation scholar and was supported by the National Science Foundation Graduate Research Fellowship Program under Grant No. DGE-1762114. TB is an Investigator of the Howard Hughes Medical Institute. This project was supported by funds from the HHMI COVID-19 Collaboration Initiative awarded to the Fred Hutchinson Cancer Research Center and the University of Washington. We thank data generators in the United States for generously sharing SARS-CoV-2 sequence data to open databases. Without open data sharing, this work would not be possible. We also thank a previous anonymous reviewer for multiple suggestions.

## Supplemental Appendix

**Figure S1.**
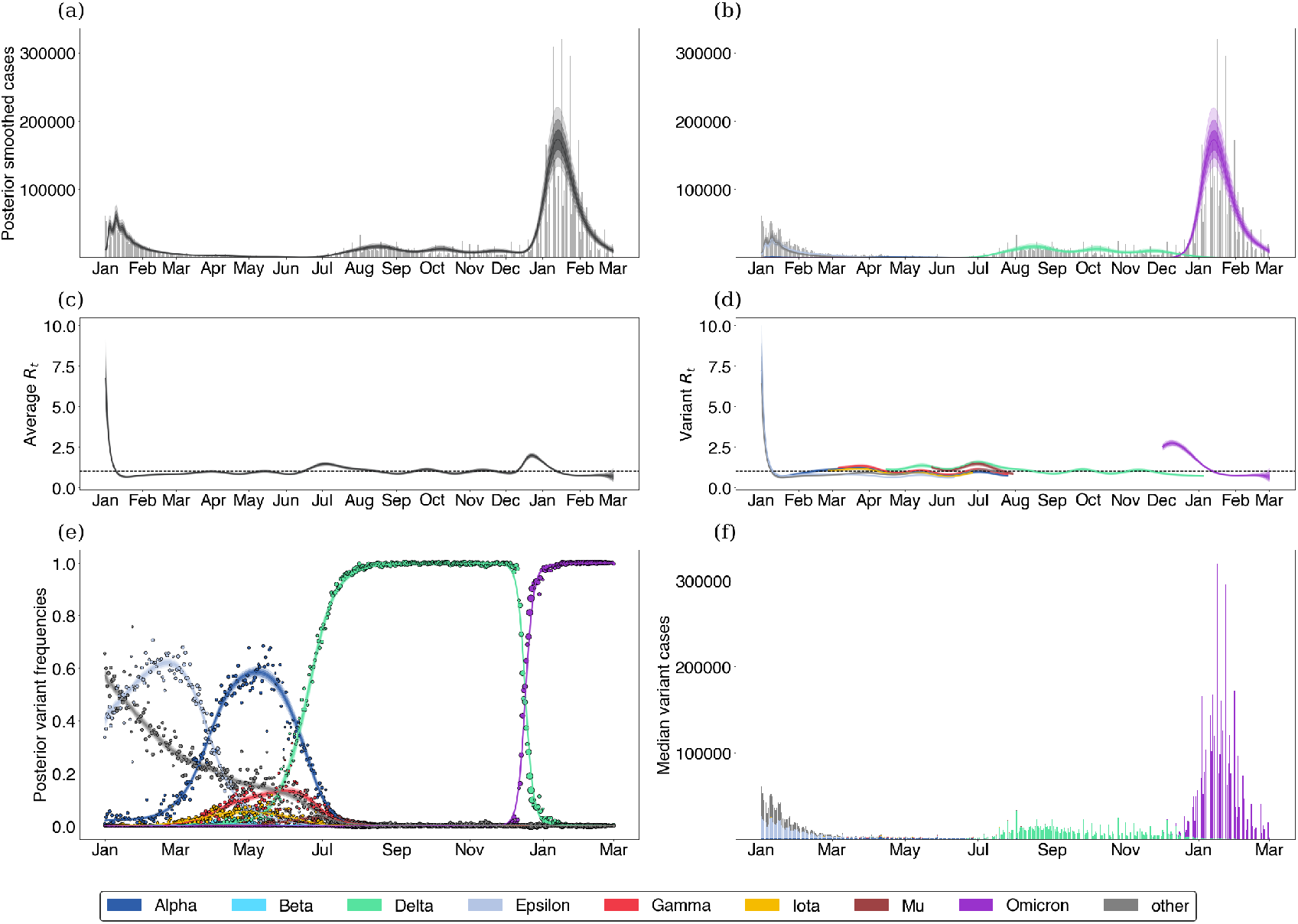
Fitting the GARW model to California data.

**Figure S2.**
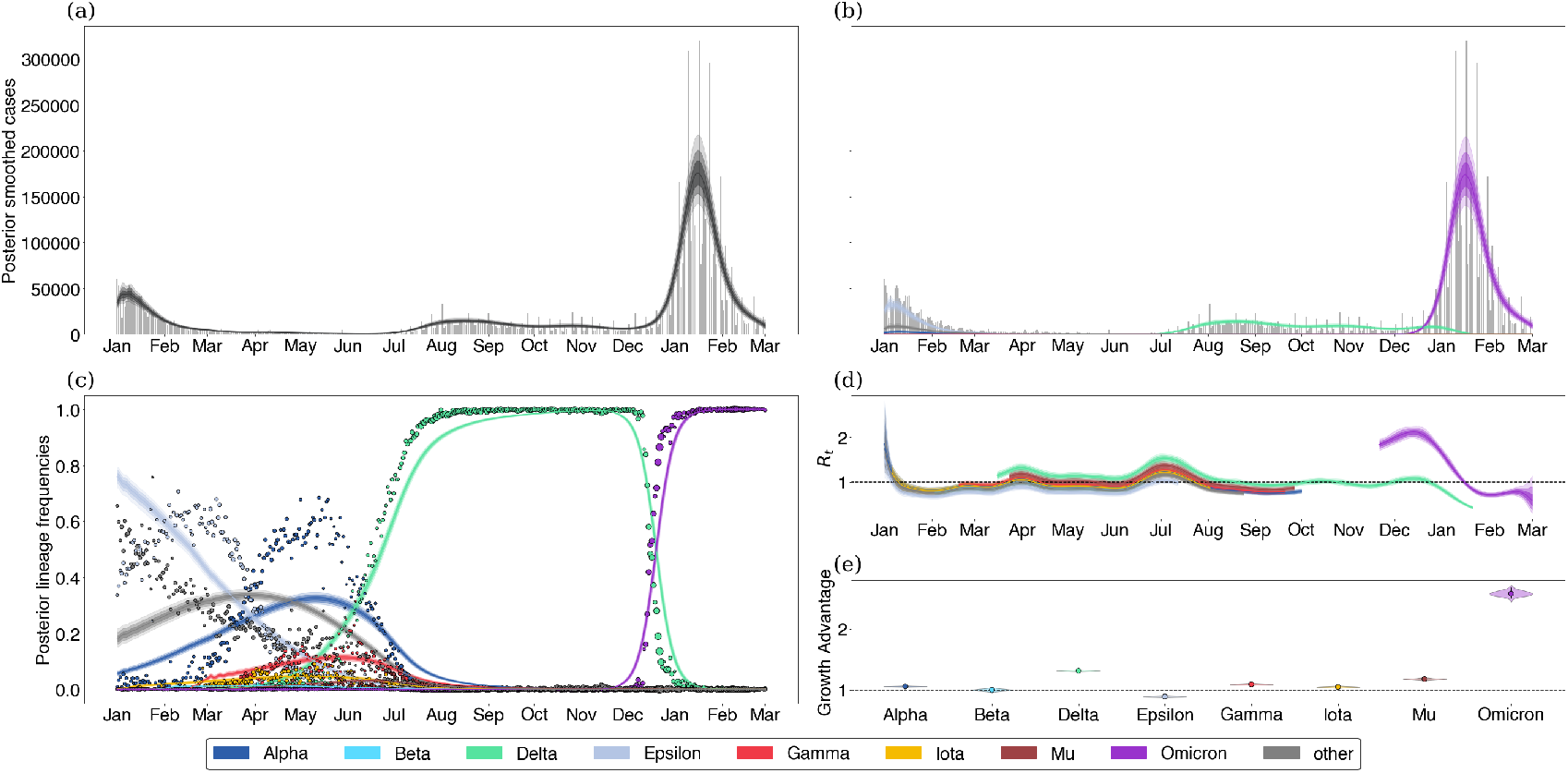
Fitting the fixed growth advantage model to California data.

**Figure S3.**
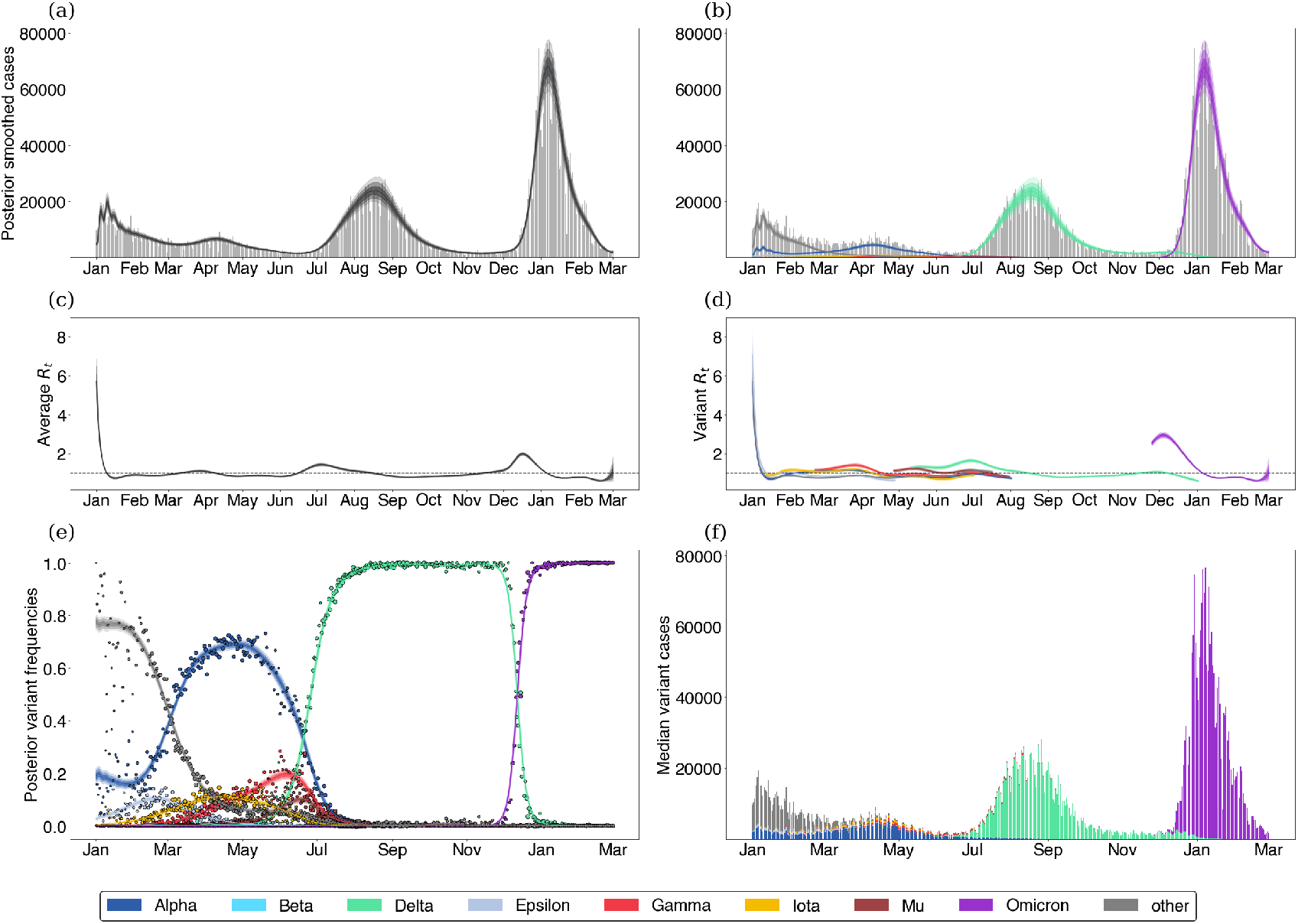
Fitting the GARW model to Florida data.

**Figure S4.**
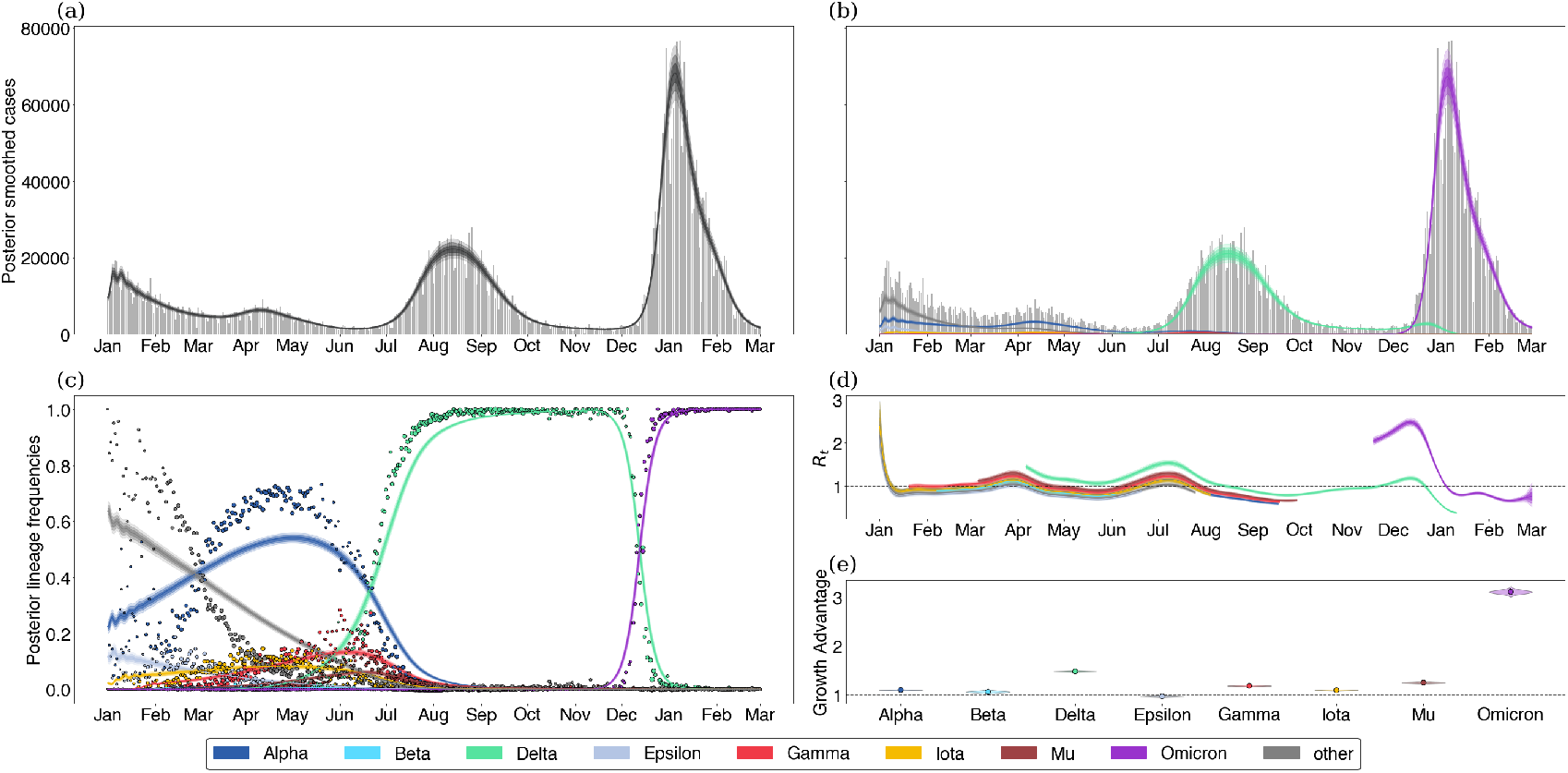
Fitting the fixed growth advantage model to Florida data.

**Figure S5.**
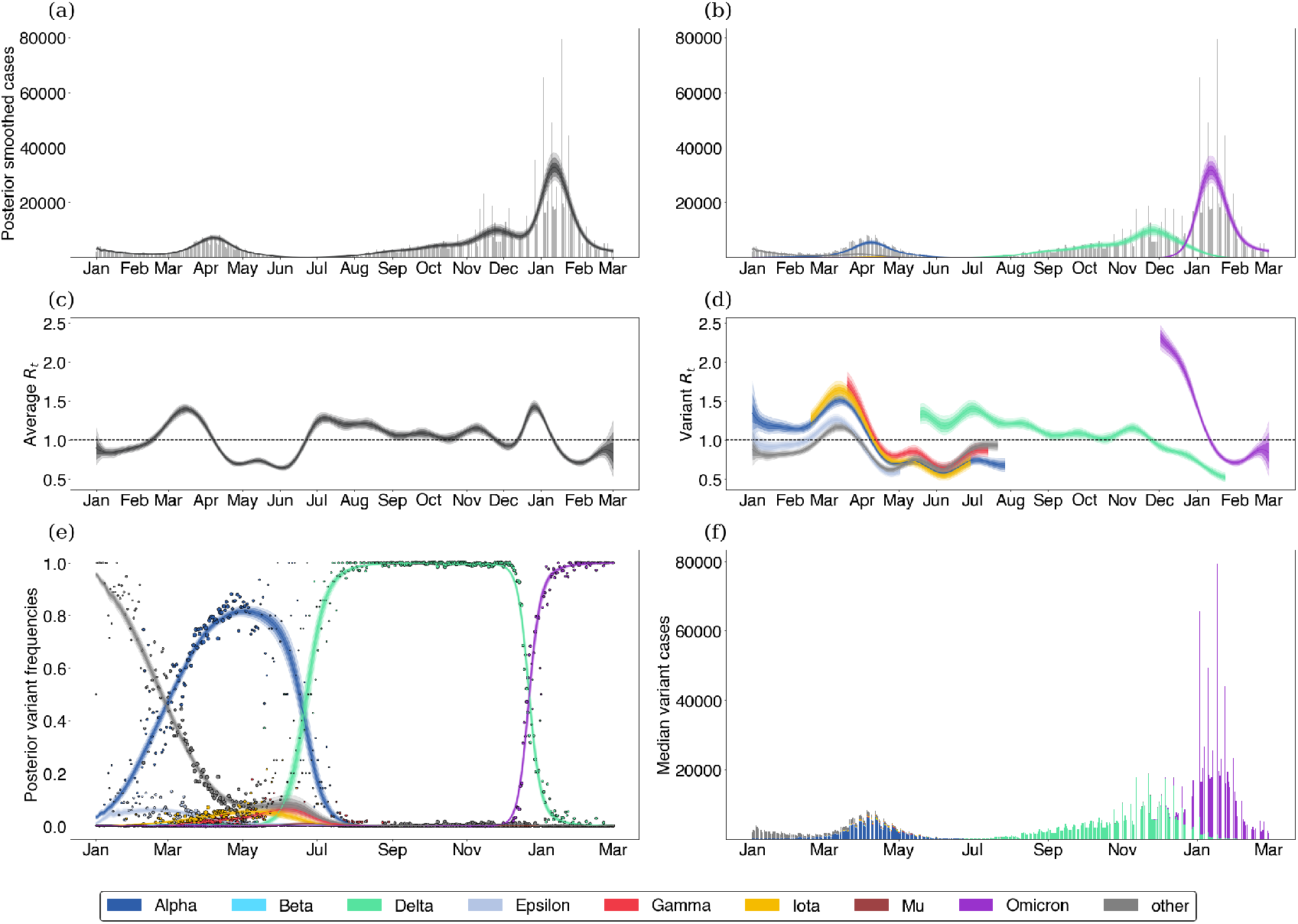
Fitting the GARW model to Michigan data.

**Figure S6.**
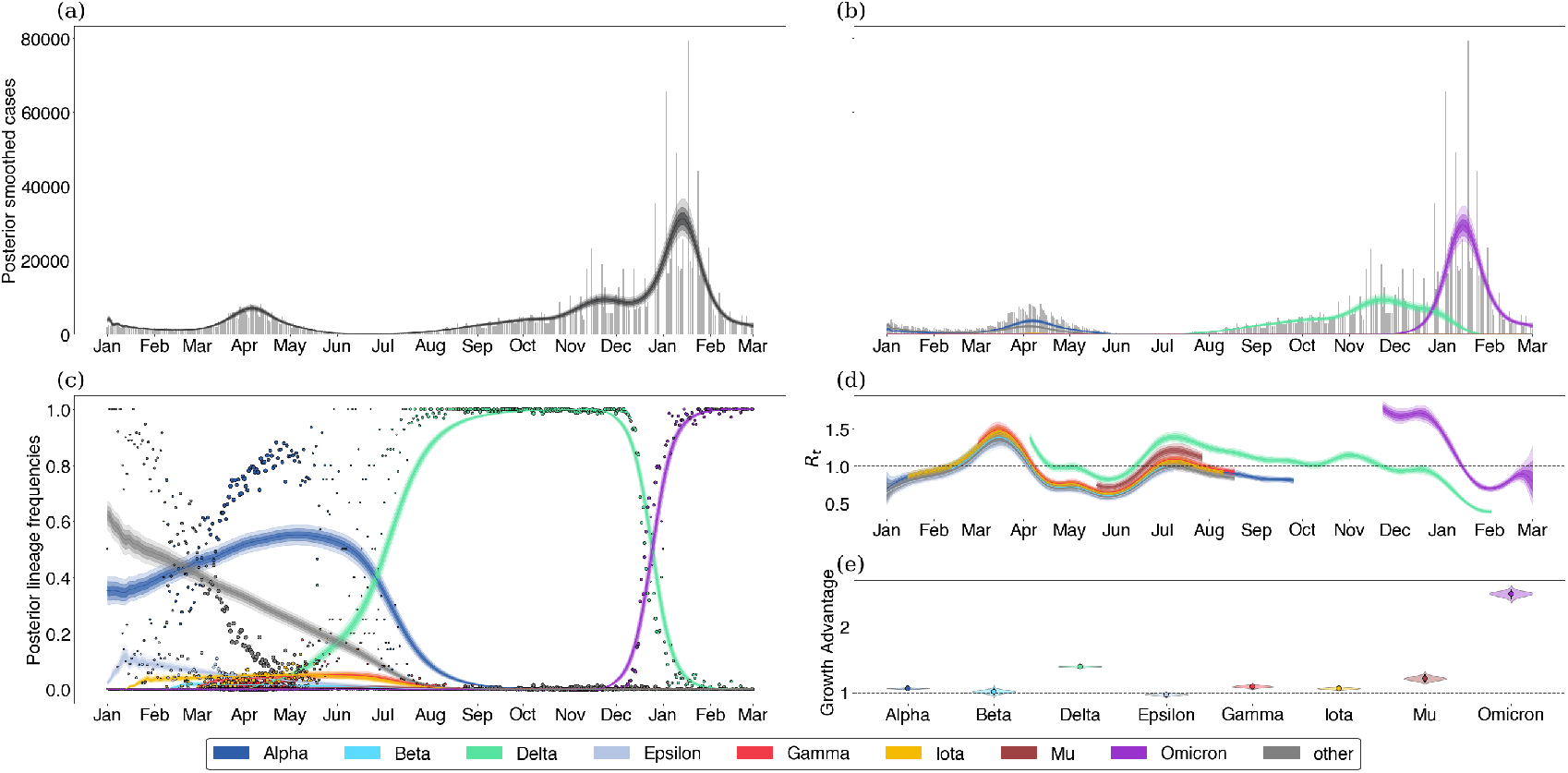
Fitting the fixed growth advantage model to Michigan data.

**Figure S7.**
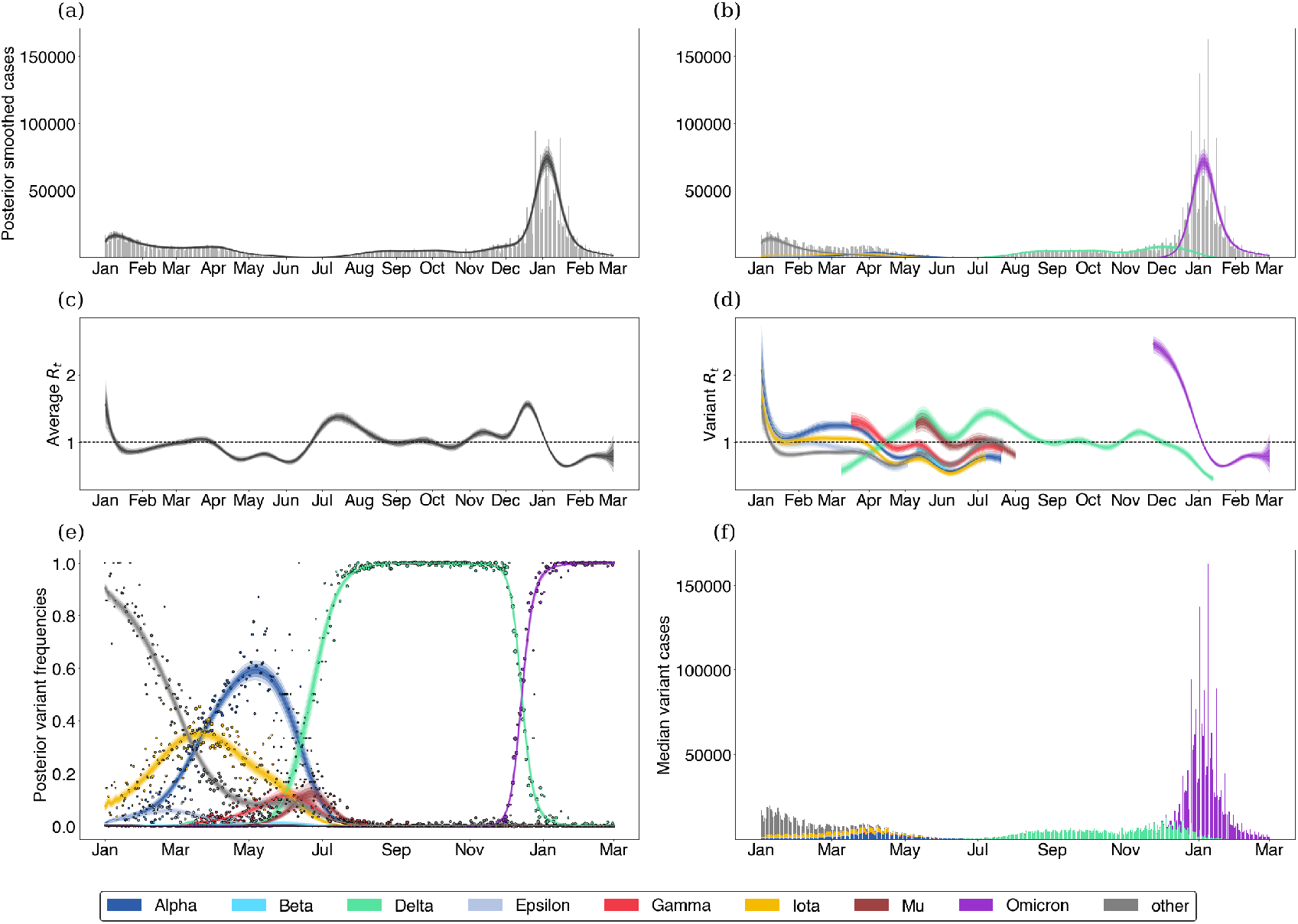
Fitting the GARW model to New York state data.

**Figure S8.**
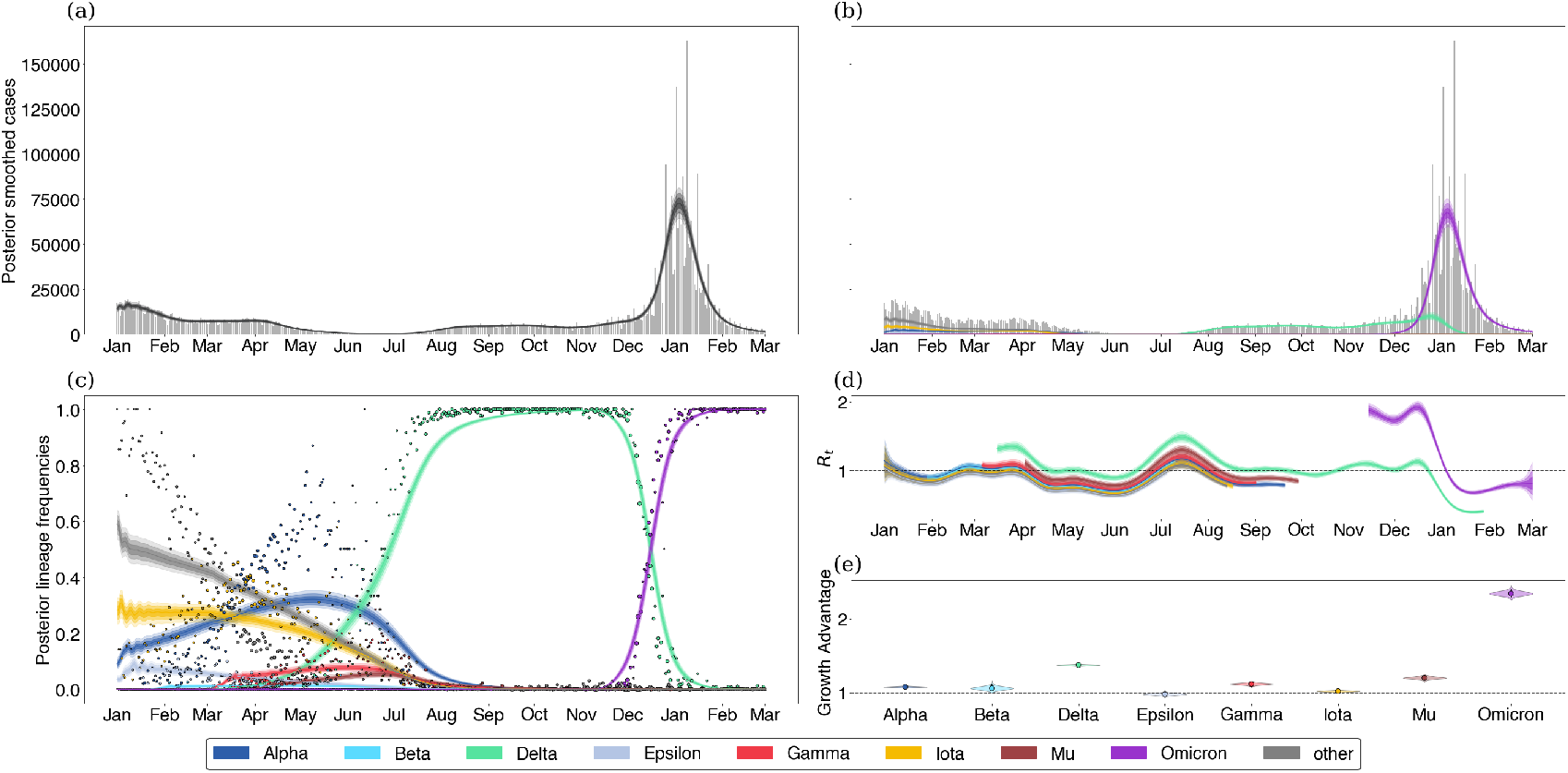
Fitting the fixed growth advantage model to New York state data.

## Supplemental Results

### Relationship to multinomial logistic regression

Other papers have tried to infer growth advantages of variants from sequence data alone, we show that the multinomial logistic regression model typically used in these analysis is roughly equivalent to our fixed growth advantage model, but that inferring relative effective reproduction numbers between variants using multinomial logistic regression requires additional restrictions on the generation time. Multinomial logistic regression typically models the probability of a given observation belong to class *v* at time *t* as

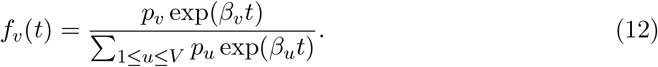

For our purpose, we can assume this probability is equivalent to the true frequency of variant *v* in the population and in this case, *p*_*v*_ is considered to be related to the prevalence on variant *v* in the population at *t* = 0 and *β*_*v*_ can be considered to be the growth advantage relative to a pivot class *u*_*∼*_ which has 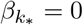. In order to see the connection between the above model and ours, we return to the original renewal equation of the form

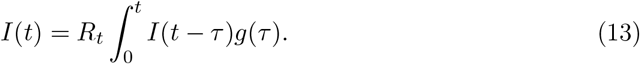

Assuming that *g* is a point mass at a mean generation time *T*_*g*_, we have that

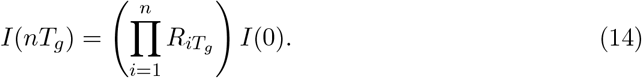

Assuming that there are several variants following these same dynamics, we have that the frequency of a given variant *v* can be written as

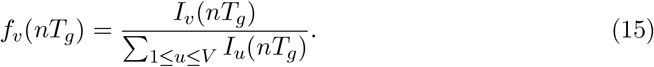

If we assume a constant growth advantage as in our model, we then have that *R*_*t,v*_ = Δ_*v*_*R*_*t*_, so that

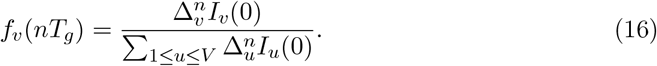

Writing Δ_*v*_ = exp(*δ*_*v*_) and *t* = *nT*_*g*_, allows us to see that

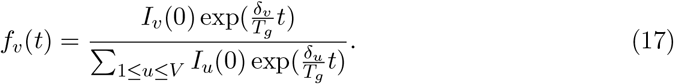

By fixing one pivot class so that 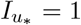 and 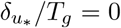, we can identify our model with the multinomial logistic regression by relating the parameters as

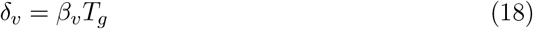

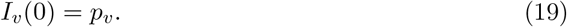

**Figure S9.**
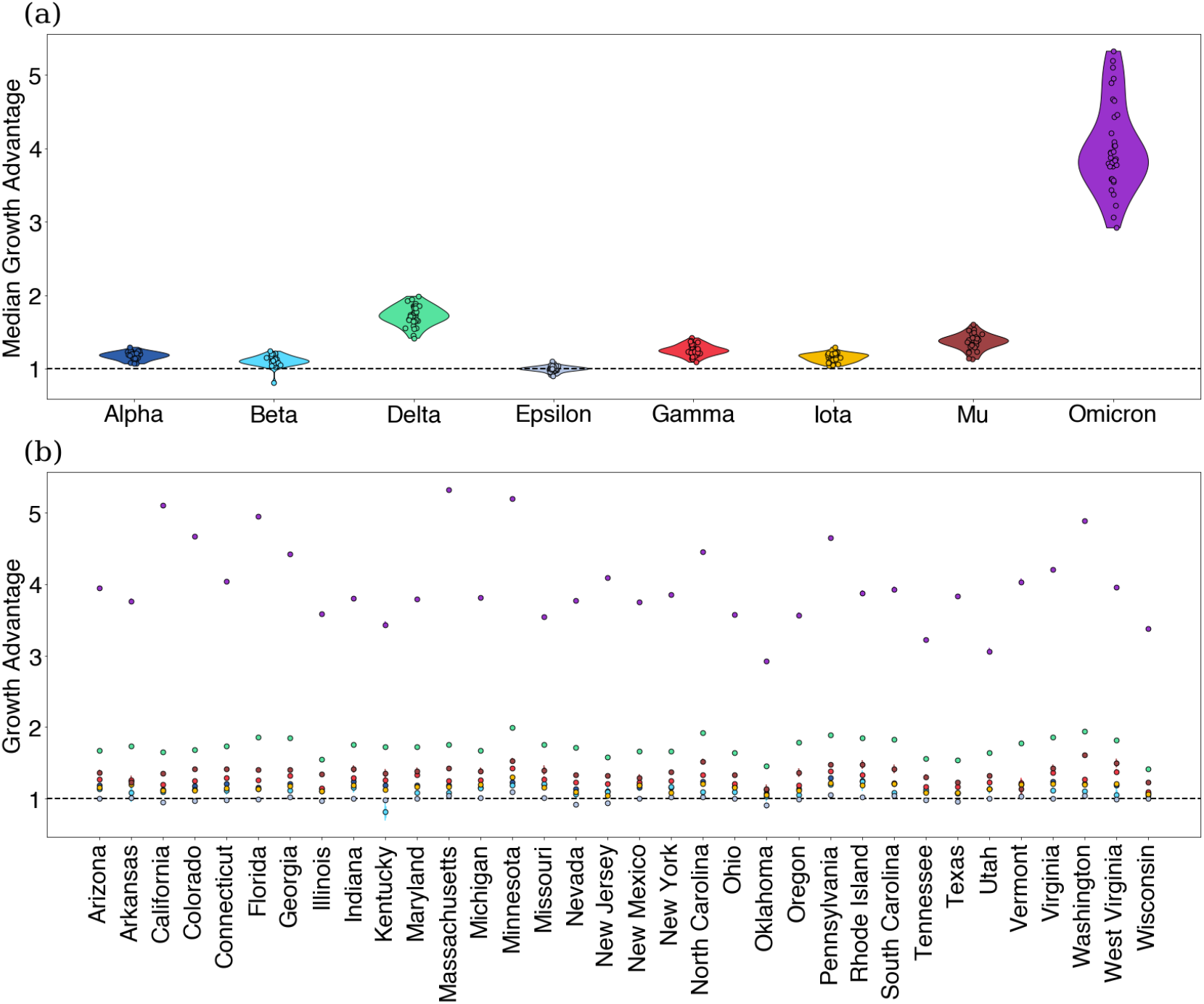
Estimating variant growth advantages in various states using Multinomial Logistic Regression model assuming generation time *T*_*g*_ = 5.2. (a) Growth advantages visualized by state. (b) Same as (a) but grouped by variant.

This shows that the multinomial logistic regression functions similarly to our fixed growth advantage model except with the additional assumption that the generation time is a point mass at *T*_*g*_. This assumption additionally allows us to relate the epidemic growth rate *r* and the effective reproduction number as *R* = exp(*rT*_*g*_) [36]. Therefore, by further assuming that the variant infections are exponentially growing with rates *r*_*v*_, we can then identify 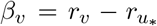. This means that the relative effective reproduction number for any two variants can be written as

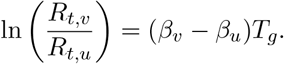

### Relating epidemic growth rates to relative effective reproduction numbers

An important relationship of interest is between the epidemic growth rate of an epidemic and its effective reproduction number. In the case of our analysis, we are particularly interested in the effect of generation time assumptions on estimated variant-specific effective reproduction numbers. First, notice that the effective reproduction number and the epidemic growth rate of an epidemic are related by

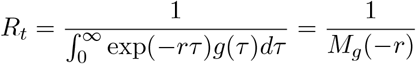

according to the Lotka-Euler equation [36] where *r* is the epidemic growth rate and *M*_*g*_ is the moment-generating function of the generation time *g*.

This allows us to write the relative reproduction number of two variants *v* and *u* as a function of their epidemic growth rates, so that

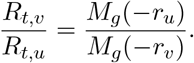

We’ll consider three common generation time assumptions. First, we consider the case where the generation time is a point mass at *T*_*g*_. In which case, *M*_*g*_(−*r*) = exp(−*rT*_*g*_) and we recover the relationship

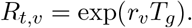

In this case, the relative effective reproduction number will depend on only the difference between the epidemic growth rates and therefore, is commonly used when converting estimated growth advantages to relative reproduction numbers in the case of logistic growth models.

Second, we consider the case where the generation time is an exponential distribution with mean *T*_*g*_. This assumption is often implicit and common in models of infectious diseases such as ODEs and their stochastic variants. Using the corresponding moment-generating function, we see that

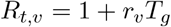

Next, we consider the Gamma distributed generation times with mean *T*_*g*_ and standard deviation *s*. This is often used in models of infectious diseases via the chain trick in which multiple compartments are chained together to obtain non-exponential generation times or infectious periods. Re-parameterizing the Gamma distribution in terms of its mean and standard deviation and using its moment generating function, we have that

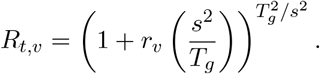

From this equation, we can see that increases in the mean of the generation time of *v* leads to decreasing estimates of *R*_*t,v*_ during epidemic decline (*r*_*v*_ *<* 0) and increased estimates during epidemic growth (*r*_*v*_ > 0) assuming *r*_*v*_ and *s* are fixed. Additionally, increases in the standard deviation will generally lead to lower inferred variant advantages. This effect is also visualized in Figure S10.

### Variant growth-advantages are sensitive to generation time

In the case where we have two variants *u, v* with Gamma-distributed generation times with means *T*_*u*_, *T*_*v*_ and standard deviations *s*_*u*_, *s*_*v*_ respectively, we can then write the relative effective reproduction number of *v* over *u* as

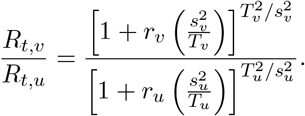

It follows that increases in the mean of the generation time of *v* leads to decreasing inferred variant advantages during epidemic decline and increased advantages during epidemic growth when all quantities are fixed. On the other hand, increases in the standard deviation will generally lead to lower inferred variant advantages.

Taking a logarithm, we can also evaluate the sensitivity of our inferred growth advantages from our fixed growth advantage model with respect to the generation time assuming it is Gamma distributed as

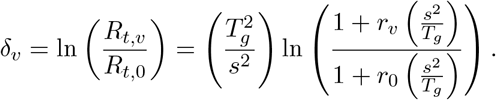

As the log of the relative effective reproduction number, the behavior here is analogous to that discussed above when the mean *T*_*g*_ and standard deviation *s* are changed. This effects of varying mean and standard deviation are illustrated in Figure S12. Although the effective reproduction number and the growth advantage appear to have strong dependence on generation time parameters, we find that the epidemic growth rate *r* is more robust to changes in generation time (see Figure S11).

The cases of exponential and Gamma-distributed generation times highlight that for non-deterministic generation times there is no guarantee that the relative effective reproduction number depends on only the difference in epidemic growth rates. In fact, these estimates based on the deterministic generation times correspond to the case in which the standard deviation shrinks zero, they are likely overestimates of variant advantages given the observed variation in the serial interval of SARS-CoV-2 infections.

### Fixed growth advantages become time-varying under generation time misspecification

We’ll now consider the case where there is a true fixed-variant growth advantage. Suppose for a two-variant system that *δ* is the constant (log) growth advantage of the variant virus over the wildtype under the variant generation time *g*_*T*_, so that 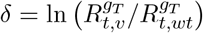. Here subscripts denote the generation time used when computing *R*_*t*_.

Under the misspecified variant generation time *g*_*M*_, we can then write the inferred growth advantage as

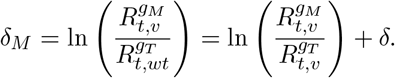

In general, the term inside the log is non-constant meaning that fixed variant growth advantages under one generation time become non-constant under generation time specification.

**Figure S10.**
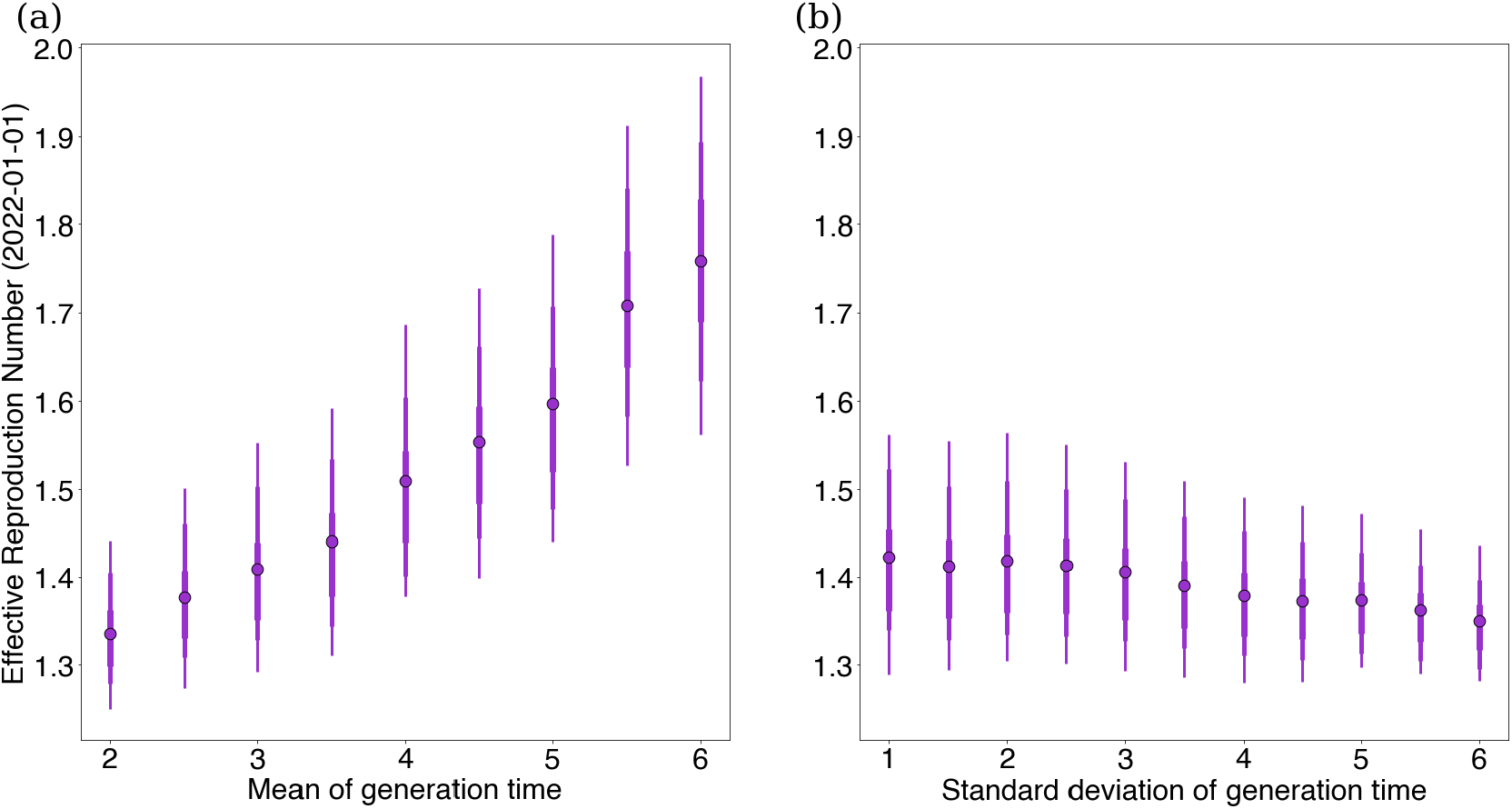
Sensitivity of effective reproduction number to changes in generation time. (a) We vary the mean of Omicron generation time keeping a constant standard deviation 1.2 and plot against effective reproduction number estimates for Omicron in Washington state on February 1st, 2022 using our GARW model. (b) The same as (a), but we instead vary the standard deviation of Omicron generation time keeping a constant mean 3.1.

**Figure S11.**
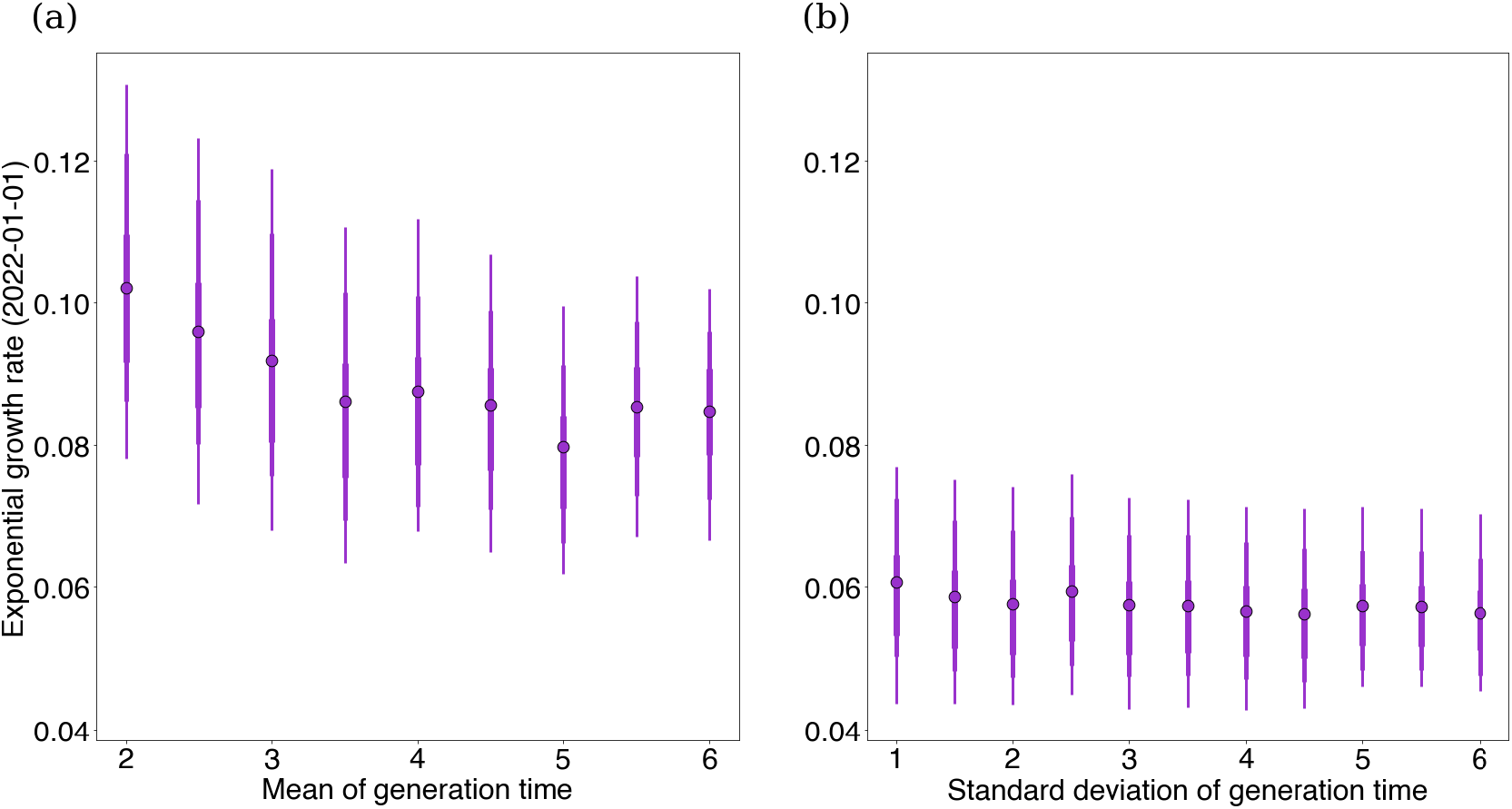
Sensitivity of epidemic growth rates to changes in generation time. (a) We vary the mean of Omicron generation time keeping a constant standard deviation 1.2 and plot against exponential growth rates for Omicron in Washington state on February 1st, 2022 using our GARW model and assuming a Gamma-distributed generation time. (b) The same as (a), but we instead vary the standard deviation of Omicron generation time keeping a constant mean 3.1.

**Figure S12.**
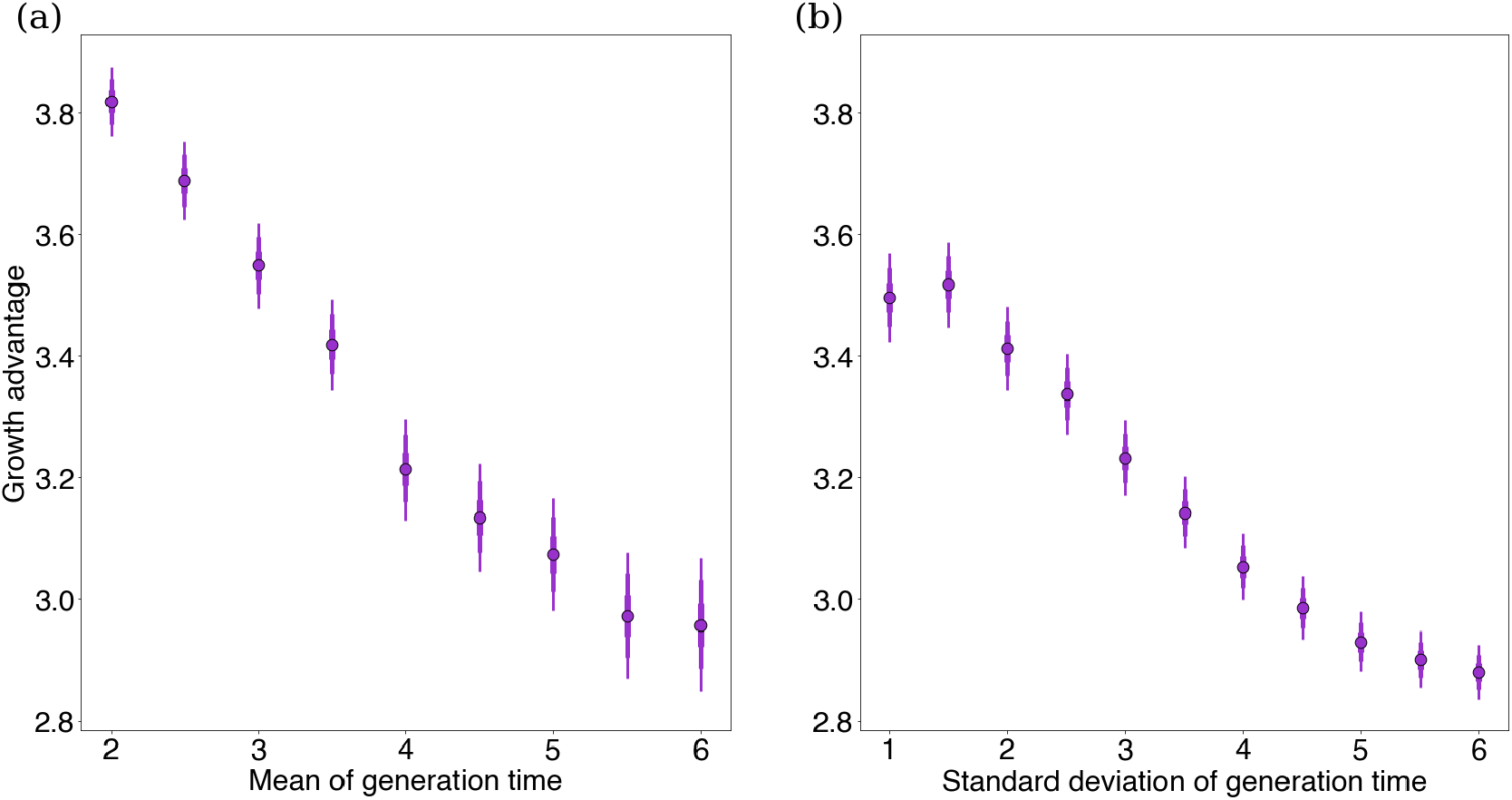
Sensitivity of growth advantages to changes in generation time. (a) We vary the mean of Omicron generation time keeping a constant standard deviation 1.2 and plot against exponential growth rates for Delta in Washington state on July 1st, 2021 using our fixed growth model. (b) The same as (a), but we instead vary the standard deviation of Omicron generation time keeping a constant mean 3.2.

